# First detection of SARS-CoV-2 Delta variant (B.1.617.2) in the wastewater of (Ahmedabad), India

**DOI:** 10.1101/2021.07.07.21260142

**Authors:** Madhvi Joshi, Manish Kumar, Vaibhav Srivastava, Dinesh Kumar, Dalipsingh Rathore, Ramesh Pandit, Chaitanya G. Joshi

**Affiliations:** Gujarat Biotechnology Research Centre (GBRC), Sector- 11, Gandhinagar, Gujarat 382 011, India; Discipline of Earth Science, Indian Institute of Technology Gandhinagar, Gujarat 382 355, India

**Keywords:** wastewater, genomic surveillance, SARS-CoV-2, mutation, health, B.1.617.2, VOCs

## Abstract

Contrary to the conventional genomic surveillance based on clinical samples (symptomatic patients), the wastewater-based genomic surveillance can identify all the variants shed by the infected individuals in the population, as it does also include RNA fragmented shredded by clinically escaped asymptomatic patients. We analyzed four samples to detect key mutations in the SARS-CoV-2 genome and track circulating variants in Ahmedabad during the first wave (Sep/ Nov 2020) and before the second wave (in Feb 2021) of COVID-19 in India. The analysis showed a total of 35 mutations in the spike protein across four samples categorized into 23 types. We noticed the presence of spike protein mutations linked to the VOC-21APR-02; B.1.617.2 lineage (Delta variant) with 57% frequency in wastewater samples of Feb 2021. The key spike protein mutations were T19R, L452R, T478K, D614G, & P681R and deletions at 22029 (6 bp), 28248 (6 bp), & 28271 (1 bp). Interestingly, these mutations were not observed in the samples of Sep and Nov 2020 but appeared before the devastating second wave of COVID-19, which started in early April 2021 in India, caused rapid transmission and deaths all over India. We found the genetic traces of the B.1.617.2 in samples of early Feb 2021 i.e., more than a month before the first clinically confirmed case of the same variant in March 2021 in Ahmedabad, Gujarat. The present study tells about the circulating variants in Ahmedabad and suggests early prediction VOCs employing the wastewater genomic surveillance approach that must be exploited at a large scale for effective COVID-19 management.

**Highlights:**

- Whole-genome sequencing of SARS-CoV-2 from the WW samples was carried out.
- Variant of Concern (VoC: VOC-21APR-02; B.1.617.2) were detected in WW samples.
- WBE may detect prevalent SARS-CoV-2 variants and monitor their cryptic transmission
- WW genomic surveillance can aid the decision-making system for public health policies.

## 1. Introduction

The deadly Severe Acute Respiratory Syndrome Corona Virus-2 (SARS-CoV-2) has a disastrous impact on human life. It continues to disrupt the public healthcare system worldwide for more than a year since the declaration of the COVID-19 pandemic by the World Health Organization (WHO) on 11 Mar 2020. SARS-CoV-2 has infected over 30 million people and caused ∼0.40 million deaths in India alone by 30 Jun 2021. The governments are taking considerable steps to expedite the vaccination drive to control the pandemic everywhere in the world. However, a public health challenge has appeared due to the high mutation rate of SARS-CoV-2 owing to its positive-sense single-stranded RNA genetic material. Mutations in the SARS-CoV-2 genome led to the emergence of different highly infectious variants of concern (VOCs). For example, the B.1.1.7 lineage of SARS-CoV-2 (VOC-20-DEC-01), which was detected in the United Kingdom (UK) in Nov 2020, is supposed to be 40-80% more contagious compared to the original strain (Davies et al., 2021; Volz et al., 2021). Likewise, other SARS-CoV-2 lineages from Brazil (P.1; VOC-21JAN-02), Southern African countries (B.1.351; VOC-20DEC-02), India (B.1.617.2; VOC-21APR-02) are more transmissible than the variants reported in early 2020. The variants of concern (VOCs) are important in terms of viral pathogenicity, virulence, and transmission. The variants of concern (VOCs) can be more transmissible, resulting in likely greater disease severity outcomes, and are also known for reduced sensitivity to antibody neutralization (Wang et al. 2021; Davies et al. 2021).

Multiple mutations in the spike protein and other important genomic areas are common in these variants, leading to attenuated efficacy of SARS-CoV-2 therapeutic interventions. For example, E484K mutation is found in the receptor binding ridge of the spike protein, which has been identified in many lineages, including B.1.351 (VOC-20DEC-02), P.1 (VOC-21JAN-02), A.23.1 (VUI-21FEB-01), B.1.525, B.1.1.318, P.2 (VUI-21JAN-01), B.1.324.1, a subclade of B.1.526, and P.3 (VUI-21MAR-02). This particular mutation reduces virus binding to polyclonal sera (Greaney et al., 2021a) and evades virus from the treatment with monoclonal antibody REGN10933, which is one of the antibodies in the REGN-COV2 cocktail (Starr et al. 2021). Mutation E484K also leads to escape from class 2 antibodies and results in a 5-fold (approx.) reduction in neutralization by COV47 plasma (Greaney et al., 2021b). Similarly, P681H and P681R mutations are present in the proximity of the furin cleavage site in the viral spike glycoprotein. P681H mutation has been reported in B.1.1.7 (VOC-20DEC-01), B.1.1.318, and P.3 (VUI-21MAR-02) lineages, while P681R mutation has been witnessed in A23.1 and all B.1.617 lineages. Both P681H and P681R mutations are supposed to enhance the cleavage of the spike protein and augment viral fusion to the host cell (Brown et al., 2021; Saito et al., 2021). Though the latter implication of P681H mutation is not clear; however, it is assumed to be responsible for the enhanced transmissibility of the B.1.1.7 variant similar to the P681R. Also, D614G mutation in spike protein is known to responsible for augmented transmissibility of the SARS-CoV-2 (Korber et al. 2020). Therefore, it is imperative to track existing circulating variants and dominant mutations in order to identify quickly developing novel variants to ensure a better decision-making system for public health policies and management of COVID-19 outbreaks.

Since it is a proven fact that COVID-19 patients excrete virus particles in the feces (Chan et al., 2020; Cheung et al., 2020); therefore RT-qPCR has been used to detect and quantify SARS-CoV-2 RNA in wastewater worldwide (Kumar et al., 2020; 2021a; 2021b; 2021c). The wastewater-based epidemiology surveillance is getting recognition worldwide due to the early prediction, large population coverage, cheaper, and more accuracy. The World Health Organization (WHO) recognized the environmental sewage surveillance strategies to monitor and detect the viral pathogens in circulation. The tracking of SARS-CoV-2 genomic variants from wastewater could also provide a better insight into their origin, pathogenicity, and transmission. However, it is challenging due to the heterogeneity and complex nature of the samples with fragmented nucleic acids. Genomic surveillance of wastewater may prove its worthiness as a powerful tool for detecting, identifying, predicting, and developing an early system for identifying VOCs in circulation to support public health interventions. There are only a few studies available in the scholarly world that attempted to sequence the SARS-CoV-2 genome and identified genomic variants from wastewater samples in different countries such as Montana, USA (Nemudryi et al., 2020), California, USA (Crits-Christoph et al., 2021), Switzerland (Jahn et al., 2021), London (Wilton et al., 2021), Canada (Landgraff et al., 2021), etc. However, it is essential to promote this novel approach at a much broader scale worldwide and develop a repository of regionally frequent mutations and dominant variants in circulation for effective management of the COVID-19 pandemic in a coordinated manner among nations.

The second wave of COVID-19 badly affected the whole country, and Gujarat was also one of the worst affected states in India, with a total ∼5 lakhs new cases and deaths of ∼5 thousand people from 1 Apr 2021 to 1 Jun 2021 (COVID 19 INDIA). Therefore, keeping in mind the present COVID-19 scenario and key role and importance of VOCs in knowing the outbreaks, transmission, epidemiology, and management of COVID-19 among populations, we performed SARS-CoV-2 genome sequencing in freshwater/ wastewater samples during the first wave and before the second wave of COVID-19 in India and compared them with the reference of Wuhan/Hu-1/2019 (EPI_ISL_402125) variant, with three prime objectives: i) detection of the existing circulating variants and dominant mutations among the populations through SARS-CoV-2 genome sequencing in freshwater/ wastewater; ii) explicate the nexus between dominant variant and the pandemic situation in Gujarat, India; iii) assess the potential of SARS-CoV-2 genomic surveillance approach in freshwater/ wastewater as an early warning indicator system to detect rapidly emerging new variants.

This is the first report of the SARS-CoV-2 whole-genome sequencing from the wastewater samples in Ahmedabad, Gujarat, aimed at the genomic mutations and identification of novel variants in the circulation.

## 2. Methodology

### 2.1 Study area and sample collection

Ahmedabad is the seventh-largest city in India and the second biggest trade center in the western Indian region, with a population of 5.5 million (Census of India, 2011). In the present study, six samples were collected, including freshwater and wastewater for analysis. Two samples were collected from the Sabarmati River in the month of Sep 2020. Likewise, two untreated wastewater samples were collected from Vinzol sewage treatment plant in Ahmedabad in Nov 2020. In Feb 2021, two samples (Untreated and treated WW) from Vinzol treatment plant were collected for analysis.

The samples were collected by grab hand sampling using 250 mL sterile bottles (Tarsons, PP Autoclavable, Wide Mouth Bottle, Cat No. 582240, India). Simultaneously, blanks in the same type of bottle were examined to know any contamination during the transport. The samples were kept cool in an ice-box until further process. The analysis was performed on the same day after bringing the samples to the laboratory. All the analyses were performed in Gujarat Biotechnology Research Centre (GBRC), a laboratory approved by the Indian Council of Medical Research (ICMR), New Delhi.

### 2.2 RNA extraction, library preparation, sequencing, and data analysis

RNA was extracted as described by the author’s earlier studies (Kumar et al., 2020; 2021a; 2021b) using the NucleoSpin® RNA Virus isolation kit (Macherey-NagelGmbH & Co. KG, Germany). The virus particles were enriched through the polyethylene glycol (PEG) method (Kumar et al., 2020; 2021a; 2021b). The extracted RNA was subjected to cDNA synthesis using SuperScript-III First-Strand Synthesis System (Invitrogen/Thermo Fisher Scientific). We used the Ion AmpliSeq Community SARS-CoV-2 panel and Ion AmpliSeq library kit Plus (Invitrogen/Thermo Fisher Scientific) for library preparation. The quality of the library was evaluated on Bioanalyzer (Agilent 2100) using DNA High Sensitivity (HS) Kit (Agilent). Further, sequencing was carried out on Ion S5 Plus System (Thermo Fisher Scientific) on 530 Chip and 400 bp chemistry.

It is noted that a single composite sample was prepared by pooling equal concentrations of extracted RNA of Sabarmati River samples (Sept 2020). Likewise, another composite sample was prepared for wastewater samples of Nov 2020. Therefore, four final samples were used for library preparation, sequencing, and data analysis (Table 1).

**Table 1.**
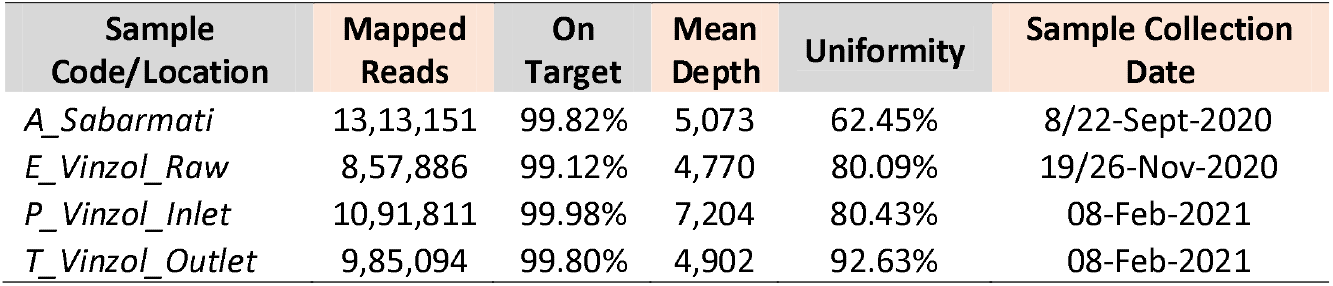
Wastewater genomic surveillance of covid-19 in Gujarat, INDIA

### 2.3 Data filtering, trimming, and genome assembly

All raw sequences were processed using the PRINSEQ-lite v.0.20.4 program for data filtering (Schmieder and Edwards, 2011). Reads were trimmed from the right where the average quality of the 5 bp window was lower than QV25, 5 bp from the left end was trimmed. Reads with lengths lower than 50 bp with average quality QV25 were removed. Quality filtered data were assembled using reference-based mapping using CLC Genomics Workbench version 12.0.3. Mapping tracks were used for variant calling and identification of the mutations. Haplotyping of the assembled genomes was carried out based on the 80% (Major allele) and 20% (Minor allele) frequency. These variants were verified and confirmed using Integrative Genomics Viewer (IGV) after manual curation. Further, Pango-Lineages were identified using the Pango-lineage classification system (https://cov-lineages.org/).

## 3. Result and discussion

The variants of concern (VOCs) are important in terms of viral pathogenicity, virulence, and transmission. Identifying the circulating variants from the wastewater/ freshwater samples could provide critical information about the possible origin, transmission, and epidemiology of SARS-CoV-2 at the local, national, and regional levels. In this backdrop, we attempted to obtain the signature of SARS-CoV-2 genomic variants from freshwater and wastewater samples during the first wave (Sep/ Nov 2020) and wastewater samples prior to the second wave (Feb 2021) of the COVID-19 in India (Table 1). This study highlights the nexus between the dominant circulating variants from freshwater/ wastewater samples and the ongoing pandemic situation in India.

The authors noticed key spike protein mutations in the SARS-CoV-2 genome assembly when compared to the reference Wuhan/Hu-1/2019 (EPI_ISL_402125) variant. The analysis showed a total of 35 mutations in the spike protein across four samples categorized into 23 types. The key mutations included Thr19Arg, Asp614Gly (D614G) in both freshwater and wastewater samples of Sep and Nov 2020 (Table 2a & 2b). Likewise, main mutations comprising C21618G/Thr19Arg (T19R), T22917G/Leu452Arg (L452R), C22995A/Thr478Lys (T478K), A23403G/Asp614Gly (D614G), and C23604G/Pro681Arg (P681R) noticed in the SARS-CoV-2 genomes from the samples collected in Feb 2021 (Table 2c & 2d). In addition, deletions at 22029 (6 bp), 28248 (6 bp), and 28271 (1 bp) were identified in wastewater samples collected in Feb 2021. These mutations in the SARS-CoV-2 genome were found similar to that of VOC-21APR-02; B.1.617.2 lineage (Delta variant).

**Table 2.**
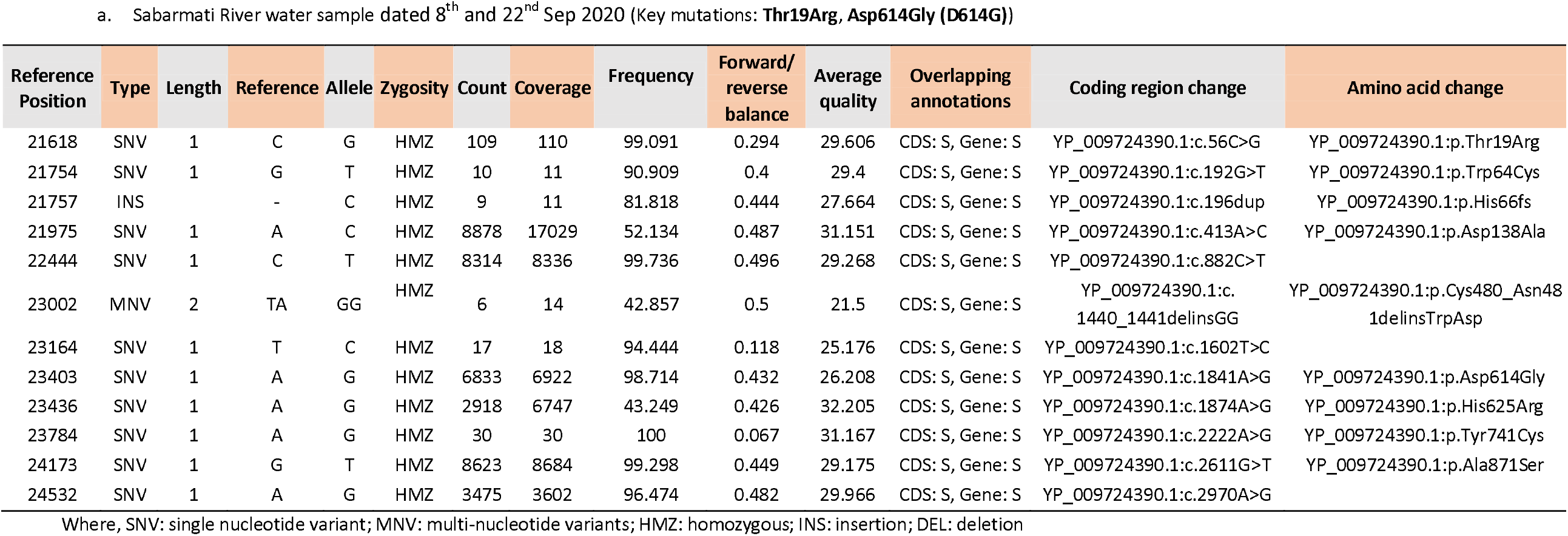
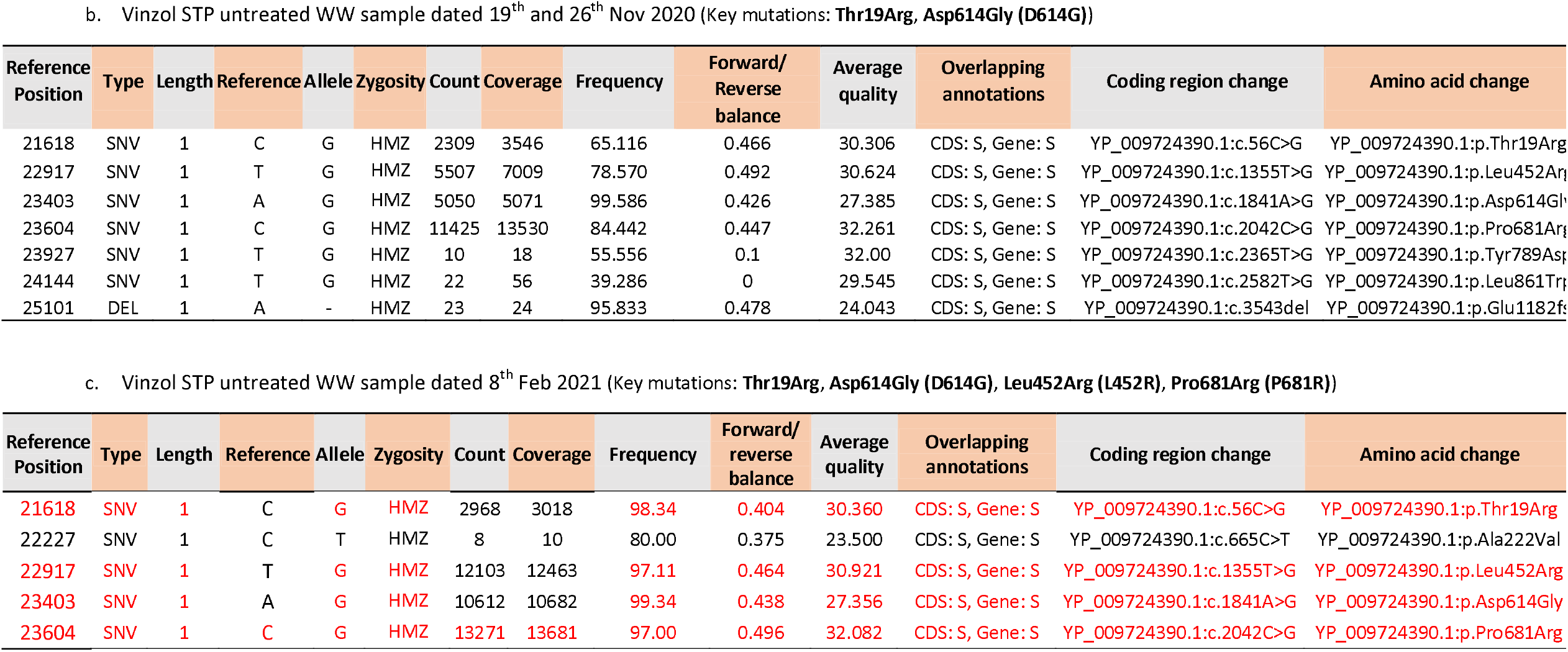
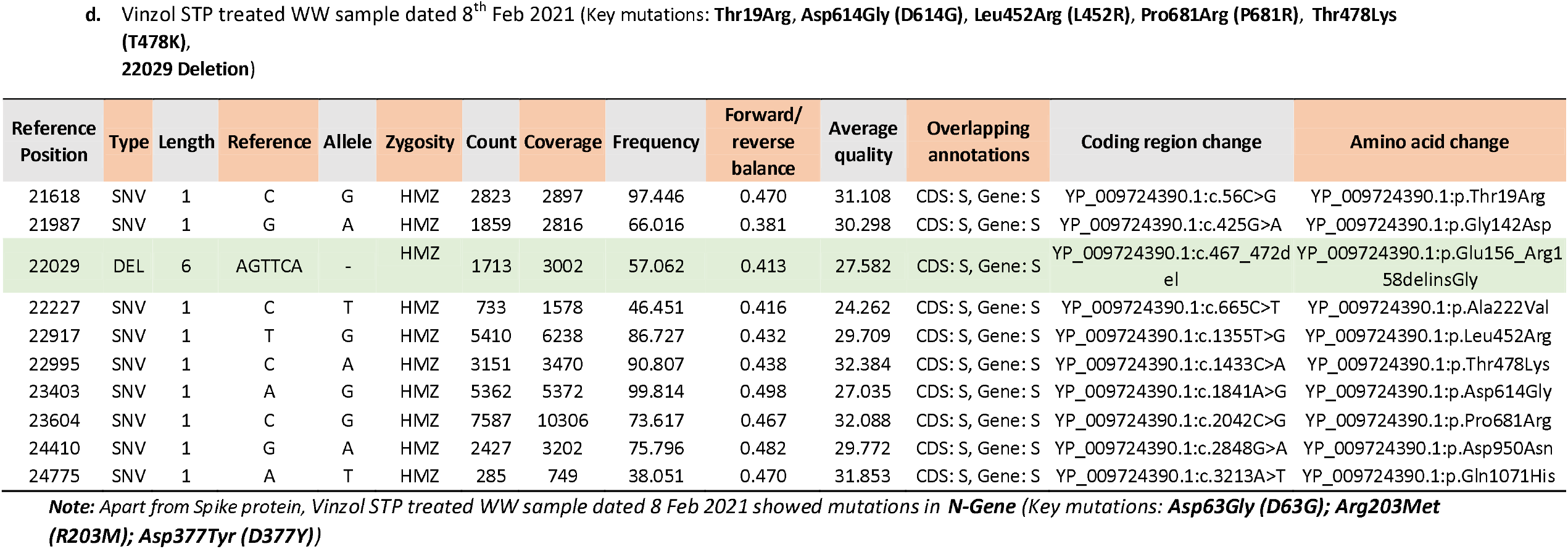
Variants of the spike protein from fresh and wastewater samples: a) Sabarmati River (water sample dated 8^th^ and 22^nd^ Sep 2020); b) Vinzol STP (untreated dated 19^th^ and 26^th^ Nov 2020); c) Vinzol STP (untreated dated 8 Feb 2021); d) Vinzol STP (treated dated 8 Feb 2021)

| Reference Position | Type | Length | Reference | Allele | Zygosity | Count | Coverage | Frequency | Forward/reverse balance | Average quality | Overlapping annotations | Coding region change | Amino acid change |
| --- | --- | --- | --- | --- | --- | --- | --- | --- | --- | --- | --- | --- | --- |
| 21618 | SNV | 1 | C | G | HMZ | 109 | 110 | 99.091 | 0.294 | 29.606 | CDS: S, Gene: S | YP_009724390.1:c.56C>G | YP_009724390.1:p.Thr19Arg |
| 21754 | SNV | 1 | G | T | HMZ | 10 | 11 | 90.909 | 0.4 | 29.4 | CDS: S, Gene: S | YP_009724390.1:c.192G>T | YP_009724390.1:p.Trp64Cys |
| 21757 | INS |  | - | C | HMZ | 9 | 11 | 81.818 | 0.444 | 27.664 | CDS: S, Gene: S | YP_009724390.1:c.196dup | YP_009724390.1:p.His66fs |
| 21975 | SNV | 1 | A | C | HMZ | 8878 | 17029 | 52.134 | 0.487 | 31.151 | CDS: S, Gene: S | YP_009724390.1:c.413A>C | YP_009724390.1:p.Asp138Ala |
| 22444 | SNV | 1 | C | T | HMZ | 8314 | 8336 | 99.736 | 0.496 | 29.268 | CDS: S, Gene: S | YP_009724390.1:c.882C>T |  |
| 23002 | MNV | 2 | TA | GG | HMZ | 6 | 14 | 42.857 | 0.5 | 21.5 | CDS: S, Gene: S | YP_009724390.1:c.1440_1441delinsGG | YP_009724390.1:p.Cys480_Asn481delinsTrpAsp |
| 23164 | SNV | 1 | T | C | HMZ | 17 | 18 | 94.444 | 0.118 | 25.176 | CDS: S, Gene: S | YP_009724390.1:c.1602T>C |  |
| 23403 | SNV | 1 | A | G | HMZ | 6833 | 6922 | 98.714 | 0.432 | 26.208 | CDS: S, Gene: S | YP_009724390.1:c.1841A>G | YP_009724390.1:p.Asp614Gly |
| 23436 | SNV | 1 | A | G | HMZ | 2918 | 6747 | 43.249 | 0.426 | 32.205 | CDS: S, Gene: S | YP_009724390.1:c.1874A>G | YP_009724390.1:p.His625Arg |
| 23784 | SNV | 1 | A | G | HMZ | 30 | 30 | 100 | 0.067 | 31.167 | CDS: S, Gene: S | YP_009724390.1:c.2222A>G | YP_009724390.1:p.Tyr741Cys |
| 24173 | SNV | 1 | G | T | HMZ | 8623 | 8684 | 99.298 | 0.449 | 29.175 | CDS: S, Gene: S | YP_009724390.1:c.2611G>T | YP_009724390.1:p.Ala871Ser |
| 24532 | SNV | 1 | A | G | HMZ | 3475 | 3602 | 96.474 | 0.482 | 29.966 | CDS: S, Gene: S | YP_009724390.1:c.2970A>G |  |

| Reference Position | Type | Length | Reference | Allele | Zygosity | Count | Coverage | Frequency | Forward/Reverse balance | Average quality | Overlapping annotations | Coding region change | Amino acid change |
| --- | --- | --- | --- | --- | --- | --- | --- | --- | --- | --- | --- | --- | --- |
| 21618 | SNV | 1 | C | G | HMZ | 2309 | 3546 | 65.116 | 0.466 | 30.306 | CDS: S, Gene: S | YP_009724390.1:c.56C>G | YP_009724390.1:p.Thr19Arg |
| 22917 | SNV | 1 | T | G | HMZ | 5507 | 7009 | 78.570 | 0.492 | 30.624 | CDS: S, Gene: S | YP_009724390.1:c.1355T>G | YP_009724390.1:p.Leu452Arg |
| 23403 | SNV | 1 | A | G | HMZ | 5050 | 5071 | 99.586 | 0.426 | 27.385 | CDS: S, Gene: S | YP_009724390.1:c.1841A>G | YP_009724390.1:p.Asp614Gly |
| 23604 | SNV | 1 | C | G | HMZ | 11425 | 13530 | 84.442 | 0.447 | 32.261 | CDS: S, Gene: S | YP_009724390.1:c.2042C>G | YP_009724390.1:p.Pro681Arg |
| 23927 | SNV | 1 | T | G | HMZ | 10 | 18 | 55.556 | 0.1 | 32.00 | CDS: S, Gene: S | YP_009724390.1:c.2365T>G | YP_009724390.1:p.Tyr789Asp |
| 24144 | SNV | 1 | T | G | HMZ | 22 | 56 | 39.286 | 0 | 29.545 | CDS: S, Gene: S | YP_009724390.1:c.2582T>G | YP_009724390.1:p.Leu861Trp |
| 25101 | DEL | 1 | A | - | HMZ | 23 | 24 | 95.833 | 0.478 | 24.043 | CDS: S, Gene: S | YP_009724390.1:c.3543del | YP_009724390.1:p.Glu1182fs |

| Reference Position | Type | Length | Reference | Allele | Zygosity | Count | Coverage | Frequency | Forward/reverse balance | Average quality | Overlapping annotations | Coding region change | Amino acid change |
| --- | --- | --- | --- | --- | --- | --- | --- | --- | --- | --- | --- | --- | --- |
| 21618 | SNV | 1 | C | G | HMZ | 2968 | 3018 | 98.34 | 0.404 | 30.360 | CDS: S, Gene: S | YP_009724390.1:c.56C>G | YP_009724390.1:p.Thr19Arg |
| 22227 | SNV | 1 | C | T | HMZ | 8 | 10 | 80.00 | 0.375 | 23.500 | CDS: S, Gene: S | YP_009724390.1:c.665C>T | YP_009724390.1:p.Ala222Val |
| 22917 | SNV | 1 | T | G | HMZ | 12103 | 12463 | 97.11 | 0.464 | 30.921 | CDS: S, Gene: S | YP_009724390.1:c.1355T>G | YP_009724390.1:p.Leu452Arg |
| 23403 | SNV | 1 | A | G | HMZ | 10612 | 10682 | 99.34 | 0.438 | 27.356 | CDS: S, Gene: S | YP_009724390.1:c.1841A>G | YP_009724390.1:p.Asp614Gly |
| 23604 | SNV | 1 | C | G | HMZ | 13271 | 13681 | 97.00 | 0.496 | 32.082 | CDS: S, Gene: S | YP_009724390.1:c.2042C>G | YP_009724390.1:p.Pro681Arg |
| 21618 | SNV | 1 | C | G | HMZ | 2823 | 2897 | 97.446 | 0.470 | 31.108 | CDS: S, Gene: S | YP_009724390.1:c.56C>G | YP_009724390.1:p.Thr19Arg |
| 21987 | SNV | 1 | G | A | HMZ | 1859 | 2816 | 66.016 | 0.381 | 30.298 | CDS: S, Gene: S | YP_009724390.1:c.425G>A | YP_009724390.1:p.Gly142Asp |
| 22029 | DEL | 6 | AGTTCA | - | HMZ | 1713 | 3002 | 57.062 | 0.413 | 27.582 | CDS: S, Gene: S | YP_009724390.1:c.467_472del | YP_009724390.1:p.Glu156_Arg158delinsGly |
| 22227 | SNV | 1 | C | T | HMZ | 733 | 1578 | 46.451 | 0.416 | 24.262 | CDS: S, Gene: S | YP_009724390.1:c.665C>T | YP_009724390.1:p.Ala222Val |
| 22917 | SNV | 1 | T | G | HMZ | 5410 | 6238 | 86.727 | 0.432 | 29.709 | CDS: S, Gene: S | YP_009724390.1:c.1355T>G | YP_009724390.1:p.Leu452Arg |
| 22995 | SNV | 1 | C | A | HMZ | 3151 | 3470 | 90.807 | 0.438 | 32.384 | CDS: S, Gene: S | YP_009724390.1:c.1433C>A | YP_009724390.1:p.Thr478Lys |
| 23403 | SNV | 1 | A | G | HMZ | 5362 | 5372 | 99.814 | 0.498 | 27.035 | CDS: S, Gene: S | YP_009724390.1:c.1841A>G | YP_009724390.1:p.Asp614Gly |
| 23604 | SNV | 1 | C | G | HMZ | 7587 | 10306 | 73.617 | 0.467 | 32.088 | CDS: S, Gene: S | YP_009724390.1:c.2042C>G | YP_009724390.1:p.Pro681Arg |
| 24410 | SNV | 1 | G | A | HMZ | 2427 | 3202 | 75.796 | 0.482 | 29.772 | CDS: S, Gene: S | YP_009724390.1:c.2848G>A | YP_009724390.1:p.Asp950Asn |
| 24775 | SNV | 1 | A | T | HMZ | 285 | 749 | 38.051 | 0.470 | 31.853 | CDS: S, Gene: S | YP_009724390.1:c.3213A>T | YP_009724390.1:p.Gln1071His |

**Note:** Apart from Spike protein, Vinzol STP treated WW sample dated 8 Feb 2021 showed mutations in **N-Gene** (Key mutations: **Asp63Gly (D63G)**; **Arg203Met** **(R203M)**; **Asp377Tyr (D377Y)**)

Interestingly, these mutations were absent in the samples analyzed during the first wave but showed their presence (in Feb 2021) just before the devastating second wave of COVID-19, which started in late March 2021 in India. It is worth mentioning that the present study revealed the genetic signs of the B.1.617.2 (Delta variant) in wastewater earlier in Feb 2021, more than a month in advance of the first case of novel B.1.617.2 variant (clinical sample) in the month of Mar 2021 in Ahmedabad, Gujarat. Our findings were comparable to those of Jahn et al. (2021), who performed deep shotgun sequencing of wastewater samples and found key mutations corresponding to the novel B.1.1.7 variant in Switzerland two weeks prior to the first case of the same variant among the population. Therefore, it is evident that genomic surveillance of wastewater can give prior information about the novel SARS-CoV-2 variants (exhibiting a high mutation rate) existing within the community even before the first clinical sample analysis. Few international studies attempted to identify SARS-CoV-2 variants from wastewater, but none of them claimed the early identification of SARS-CoV-2 VOCs in wastewater before the first case of the same variant existing within the population. For example, Nemudryi et al. (2020) identified 11 single-nucleotide variants (SNVs) in the assembled genome from wastewater samples in Bozeman, Montana (USA). These SNVs were distinct from the Wuhan-Hu-1/2019 reference sequence. Likewise, Landgraff et al. (2021) identified a near-complete SARS-CoV-2 consensus level genome sequence from untreated wastewater in Canada and reported many mutations designating the B.1.1.7 SARS-CoV-2 VOC in the sample.

Apart from the early information of VOCs in wastewater, it is important to note that we observed SARS-CoV-2 variants from the treated wastewater sample, indicating that the wastewater treatment plant (WWTP) failed to remove the virus. This finding was similar to those of Kumar et al. (2021a; 2021b), who reported SARS-CoV-2 RNA in treated wastewater samples. In addition, a high mutation rate was found in treated wastewater compared to the untreated sample in Feb 2021 (Table 2c & d). This might be because the second wave of COVID-19 in India led to the increased and overconsumption of certain drugs such as Ivermectin, Azithromycin, Remdesivir Chloroquine, Favipiravir, Hydroxycholoroquinine that finally excreted through urine and feces, ultimately found their way to the WWTPs. Due to the inadequate removal efficiency of the conventional WWTP to eliminate these drugs, their residues and transformation products remain accumulated in the treatment plant (Kumar et al., 2021c). A long residence time of wastewater in WWTP where the interaction of a heterogeneous cocktail of drug residues/ intermediates with the SARS-CoV-2 virus particles might have led to the increased mutation rate and origin of new variants.

Overall, the genomic surveillance of SARS-CoV-2 variants in wastewater samples offers the information of circulating novel variants and their cryptic transmission in advance with the following advantages:

a. It is a useful approach for detecting and identifying VOCs, VUIs, and mutations of interest within a population.
b. A continuous and large-scale time-series monitoring of wastewater can identify disease outbreaks and clustering of VOCs & VUIs, and explain their genesis, virulence, transmission, and intensity of spread within a population.
c. It can give more detailed and less biased data as it covers a broader population, whereas clinical samples only represent a subset of those who went through sequencing tests.
d. It can help in identifying regions with a greater prevalence of the virus/ variants in circulation among populations which may help in zoning the city. This data can further be used to help with non-pharmaceutical interventions (NPIs).
e. It can help in assessing the success of containment and the efficacy of NPIs
f. This approach is comparatively less time-consuming, more accurate, low budget, and less manpower requiring than large-scale clinical testing and sequencing.

## 4. Key challenges of wastewater SARS-CoV-2 genomic surveillance

a. Sample enrichment via concentration methods is required owing to the low concentration of SARS-CoV-2, and the genomes may be severely damaged and fragmented.
b. Sample collection timing and intervals are critical parameters for an optimal surveillance strategy.
c. Effect of physicochemical wastewater treatment process on the false positive and negative detection limits.
d. Primer biases and sensitivity issues remain major concerns in wastewater-based genomic surveillance of the SARS-CoV-2.
e. Poor amplification of target amplicons and partial genome coverage.
f. False negatives are more likely to observe when a sample exhibits a low frequency of mutation or variant.
g. Wastewater comprises a mixed population of VOCs & VUIs, making data interpretation difficult as the genomic connection between SNPs/Indels used to assign phylogeny and subsequently lineage is lost.

## 5. Conclusions

Genomic surveillance of wastewater enables researchers to identify recent introductions of SARS-CoV-2 lineages prior to their detection by local clinical sequencing. The monitoring and presence of SARS-CoV-2 variants in wastewater over time offer a better picture of the dominant variant, transmission, and epidemiology. In the present study, key mutations were observed in the spike protein in reference to the Wuhan/Hu-1/2019 (EPI_ISL_402125) variant. A total of 35 mutations in the spike protein across four samples were noticed, categorizing into 23 types. We noticed the presence of spike protein mutations linked to the VOC-21APR-02; B.1.617.2 lineage (Delta variant) in wastewater samples of Feb 2021. The key spike protein mutations were T19R, L452R, T478K, D614G, & P681R and deletions at 22029 (6 bp), 28248 (6 bp), & 28271 (1 bp). It is worth noting that mutations which define the Delta variant were completely absent in samples during the first wave but appeared in the wastewater samples more than a month before the second wave in India and its first reported case in Ahmedabad, Gujarat. Furthermore, a greater frequency of mutation and variants was found in treated wastewater samples. Due to the long residence time of wastewater in WWTP, and interaction of heterogeneous drug with the SARS-CoV-2 virus particles may have led to the increased mutation rate and genesis of new variants. The study concludes that this approach is not only beneficial for detecting and identifying VOCs, VUIs, transmission, and epidemiology of SARS-CoV-2 but also aids in assuring adequate and resilient public health responses.

## Data Availability

All data are included in the manuscript.

## Notes

The authors declare no competing financial interest.

## Acknowledgement

This work is funded by UNICEF, Gujarat. We acknowledge the help received from GPCB and AMC.

## Notes

### Competing Interest Statement

The authors have declared no competing interest.

### Funding Statement

UNICEF- India

### Author Declarations

No such permission is required as no human subject was involved.

